# Perceived risk for hypertension and associated factors among adult persons living with HIV in Kamuli district, Uganda

**DOI:** 10.1101/2024.07.02.24309844

**Authors:** Thomas Buyinza, Freddie Ssengooba, Roy William Mayega, Joan Nankya Mutyoba, Shamim Warugaba Birungi, Geofrey Mutole, Rawlance Ndejjo

## Abstract

**Background:** Persons living with Human Immunodeficiency Virus (PLHIV) are at a higher risk of developing hypertension. Preventive measures can be taken to lower this risk, but it is important for PLHIV to be aware of this risk. However, it is unclear whether PLHIV perceive themselves to be at risk for hypertension, and if they do, it is unknown to what extent they perceive this risk. This information is crucial in designing and implementing appropriate preventive measures for hypertension targeting this high-risk population. Our study aimed to provide evidence on the level of risk perception for hypertension and associated factors among PLHIV in Kamuli District.

**Methodology:** This was a cross-sectional study conducted among 392 adult PLHIV in Kamuli District. The participants were selected using consecutive sampling and pretested structured questionnaires were used to collect data which later analysed in STATA version 15.0. Descriptive analysis was carried out and data was summarized using frequencies and means. Modified Poisson regression was used to determine factors independently associated with low-risk perception for hypertension.

**Result:** More than half, 68.1% (267/392) perceived themselves to be at low-risk for hypertension. Factors associated with low-risk perception were: residing in peri-urban area (APR=1.23; 95%CI: 1.04 −1.46) and urban areas (APR=0.73; 95%CI:0.54-0.99); having low knowledge of hypertension (APR=1.98; 95%CI: 1.55-2.53); low trust for health workers (APR=1.13; 95%CI: 1.01-1.25); having no other NCD (APR=1.38; 95%CI: 1.00-1.89); having no family history of hypertension (APR=1.42; 95%CI: 1.20-1.68); and consumption of sufficient fruits/vegetables (APR=1.13; 95%CI: 1.01-1.27).

**Recommendation:** There is need to implement appropriate health education programs specifically tailored to increasing knowledge and risk perception for hypertension among PLHIV. Health workers can integrate routine risk assessments into HIV care to identify PLHIV who are at very high-risk of developing hypertension and provide them with personalized care. This can influence adoption of hypertension preventive measures.

## Background

Hypertension is a major risk factor for non-communicable Diseases (NCDs), especially cardiovascular diseases (CVDs). Globally, 13% of all premature deaths annually are due to hypertension (HTN) [1], and by 2025, about 1.56 billion adults will be living with HTN [2]. In 2017, over 25% of persons living with HIV (PLHIV) globally were reported to be hypertensive [3]. In Uganda, Antiretroviral Therapy (ART) for PLHIV was rolled out in 2000 and since then, the morbidity and mortality rate from HIV/AIDS and related complications has been on a decline [4] accompanied by an increase in chronic degenerative diseases associated with ageing such as HTN. The dual burden of NCDs especially HTN and HIV/AIDS in Uganda and sub-Saharan Africa (SSA) in general, is of great public health concern.

PLHIV have a 1.5 to 2 times relative risk of developing CVDs at a lower median age when compared with uninfected people [5]. In Uganda, the prevalence of HTN among PLHIV is on an increasing trend, in the range of 11-29% according to recent studies in Uganda [6,7]. Specific ART regimes especially those containing Protease Inhibitors (PI) have been linked to increase in HTN, CVDs and other NCDs. Kazooba, et al. [8] reported that PI containing ART regimens were independently associated with 2-fold risks of having abnormal total cholesterol, LDL, triglycerides, low HDL and HTN compared to non-PI ART regimen. Possible mechanisms that contribute to the higher burden of HTN among PLHIV include chronic inflammation, adaptive immune activation and increased microbial translocation caused by HIV, blood vessel damage resulting from long-term ART exposure especially Highly Active ART (HAART) [9] and other risky behaviours such as excessive alcohol intake, inadequate physical activity and unhealthy dietary lifestyles [10].

Given the high burden of HTN experienced by PLHIV, consistent engagement in preventive behaviours such as adequate physical activity, healthy eating and reduced alcohol intake is critical to prevent the condition [11]. For a number of chronic diseases including HTN, symptoms do not manifest until the disease progresses [12], which influence individual’s risk perception. Disease risk perception refers to a person’s subjective judgment about the likelihood of occurrence of a negative or harmful event such as, illness, injury, extreme stress, disease and death [13]. Risk perception is an important factor for consideration in public health and risk communication because it influences which types of risk people care about and how they deal with them. Low risk perceptions can contribute to undesirable health consequences by acting as a barrier to adoption and maintenance of preventative health behaviours [14]. Perception of being at higher risk influences behavioural change resulting into higher uptake of the HTN preventive measures [15]. There is a dearth of information on levels of risk perception for HTN among PLHIV in Uganda/SSA. This study assessed the level of risk perception for HTN and the associated factors among adult PLHIV in Kamuli District.

## Methods

### Study design and setting

This was a cross sectional study based on constructs from the Health Belief Model [15] and it was conducted in public Health Centres (HCs) in Kamuli District. Kamuli is located in the Busoga region where HIV prevalence is estimated at 4.5% according to the Uganda Population-based HIV Impact Assessment report, 2020. The district has 14 public HC IIIs which provide HIV counselling and testing services, ARVs and TB screening, two HC IVs and Kamuli General Hospital which provides HTS, ARVs, viral load testing, TB and cancer screening. The study was conducted at HC III, IV plus the General Hospital because these facilities provide regular HIV care services. HIV clinics operate 2 days per week at HC III and 4 days at HC IV and the hospital. At each of the health facility, HIV care services are stratified into children, youths, maternity and adults to make them more friendly to the specific age groups.

### Study population, sample size and sampling

The study population were adult PLHIV and all were eligible for inclusion except those who reported to the health facilities for TB treatment, referred for hospitalization or need urgent care were excluded. Kish Leslie formula [16] for cross sectional studies, was used to estimate sample size of 384 based on the following assumptions: a 50% proportion of PLHIV who perceive low-risk for hypertension and a precision of 5%. Considering a non-response rate of 10%, the sample size was increased to 422. By purposive sampling, seven out of the 14 HC IIIs were selected for inclusion preferring those serving bigger number of PLHIV. Besides, the two HC IVs and Kamuli General Hospital were included. For each of the selected health facilities an estimated total number of PLHIV served per month was obtained from the HC in-charge and participants were sampled from each of the HC proportional to size. Participants were recruited into the study by consecutive sampling until the required sample size at a given health facility was obtained. Participants who self-reported as having hypertension were discontinued from the study (no longer at risk for hypertension) and they were reported as non-response. This reduced the denominator from 422 to 392 but still met our sample size of 384.

### Data collection and quality assurance

A structured questionnaire administered by Research Assistants was used for data collection, which started on 15^th^ September 2022 and ended on 30^th^ September 2022. Prior to data collection, Research assistants were trained and sensitized on the purpose of the research, appropriate procedures of informing potential participants about the research and seeking consent as well as the data collection. Besides, the Research Assistants could speak fluent English and Lusoga (the local language) to easily capture the most intended data. The data collection tool (questionnaire) was pretested among PLHIV at a neighboring Buwenge HC IV in Jinja District in the same Busoga sub region. After pretesting, the tool was reviewed, and necessary corrections done. In addition, the tool was submitted to expert NCD Researchers who gave recommendations for edition and made final approval. The approved questionnaire was used by the trained Research Assistants to collect data through direct face-to-face interviews (one-on-one).

### Measurements

The level of risk perception for HTN was assessed by adapting three measures previously validated in Uganda through a study that assessed Hepatitis B virus perceptions among pregnant women, where Cronbach’s Alpha coefficient was used to assess internal consistency of scales measuring risk perception [17]. The measures of risk perception included: - absolute lifetime risk perception, conditional risk perception and comparative risk perception. For all the three measures of risk perception, responses were on a 4-point Likert-scale and overall risk perception was obtained by dichotomizing risk perception into low-risk and high-risk perception. Responses “very low/much lower” and “low/lower” responses were combined and the new category named “low” while “high/higher” and “very high/much higher” responses were combined, naming the new category as “high.” Factors associated with the level of risk perception for HTN were assessed including the following: Level of knowledge of HTN was measured using 8 questions. Every correct response on knowledge areas was scored 1, and every incorrect response was scored 0. Bloom’s cut-off points were used to categorize knowledge ie., ≥80% score as high knowledge, 60-79% as moderate and <60% as low knowledge. Knowledge level was dichotomized by combining levels “moderate” and “high”, naming the new category as “high” Knowledge. Perceived trust in health workers was measured using items adapted from a scale that assesses trust in community health workers. The scale was developed and validated in Bangladesh, Haiti, and Kenya [18]. Five question items were used to measure trust on a 4-point Likert scale which was afterwards dichotomized by combining responses “never” and “some of the time”, naming the new category as “low trust”, and then a combination of “most of the time” and “all of the time”, naming the new category as “high trust.” Self-reported clinical factors assessed included having NCD comorbidities (defined as having experienced at least two of HTN, diabetes, cardiovascular diseases, cancer), family history of HTN (defined as having one biological family member who has ever HTN), time since HIV diagnosis, ART exposure status (naive or on ART) and duration on ART. Participants were asked to self-report each of these factors if they had or never had any of them. General body health was measured using a single item “How would you rate your current health status on a scale from 1 to 5?” where 1 was considered poorest and 5 - very good [19].

Exposure to behavioural risk factors of HTN such as excessive alcohol intake, smoking cigarette and insufficient consumption of fruits/vegetables were measured using question items adapted from the WHO stepwise approach to chronic disease surveillance [20].

### Data management and analysis

The collected data underwent cross-checking and review for completeness and in case missing data was found, a call back to the participant was made to complete the data. The data was entered into Microsoft excel and later exported to STATA version 15.0 for analysis. Descriptive analysis was carried out to provide frequency and percentage distribution of categorical variables such as sex, marital status, education level, occupation, level of knowledge and risk perception for HTN. Means and standard deviation were used to summarise continuous variables such as age, monthly income, time since HIV diagnosis, duration on ART, number of servings of fruits and/vegetables. The continuous variables were later categorized, for example. age categorized into 18-30, 31-50 and above 50 years, monthly income categorized into 0-65, 66-105 and above 105 US dollars, fruits and or vegetables consumption categorized into insufficient (if less five servings per day) and sufficient (if equal or greater than five servings per week).

Modified poisson regression was used to conduct bivariate analysis to assess factors associated with low-risk risk perception, considering a p-value <0.2. Multicollinearity was checked and sex was found to be highly correlated with age (correlation coefficient ≥0.4), thus it (sex) was dropped in the final multivariate analysis. Factors with p-value <0.2 at bivariate level of analysis were included into multivariable analysis to assess factors independently associated with low-risk perception for hypertension. Using modified Poisson regression [21], stepwise elimination procedure was used during model building, which involved adding qualified variables one at a time into multivariable analysis while dropping those with least significance. A model with a smaller Akaike’s information criterion and a log-likelihood ratio closer to zero was selected as the best fit for the data. Factors with p-value <0.05 were reported as those independently associated with low-risk perception for hypertension.

### Ethical considerations

This study was approved by Makerere University School of Public health’s Higher Degrees,Research and Ethics Committee (study protocol number 090) and each participants gave written informed consent prior to participation.

## Results

### Socio-demographic characteristics of respondents

More than half, 60.2.0% (236/392) of the respondents were females, mean age was 37.1(±14.1) years, 58.2% had not gone beyond primary level of education and 79.1% had a monthly income not exceeding 65% dollars (Table 1).

**Table 1:**
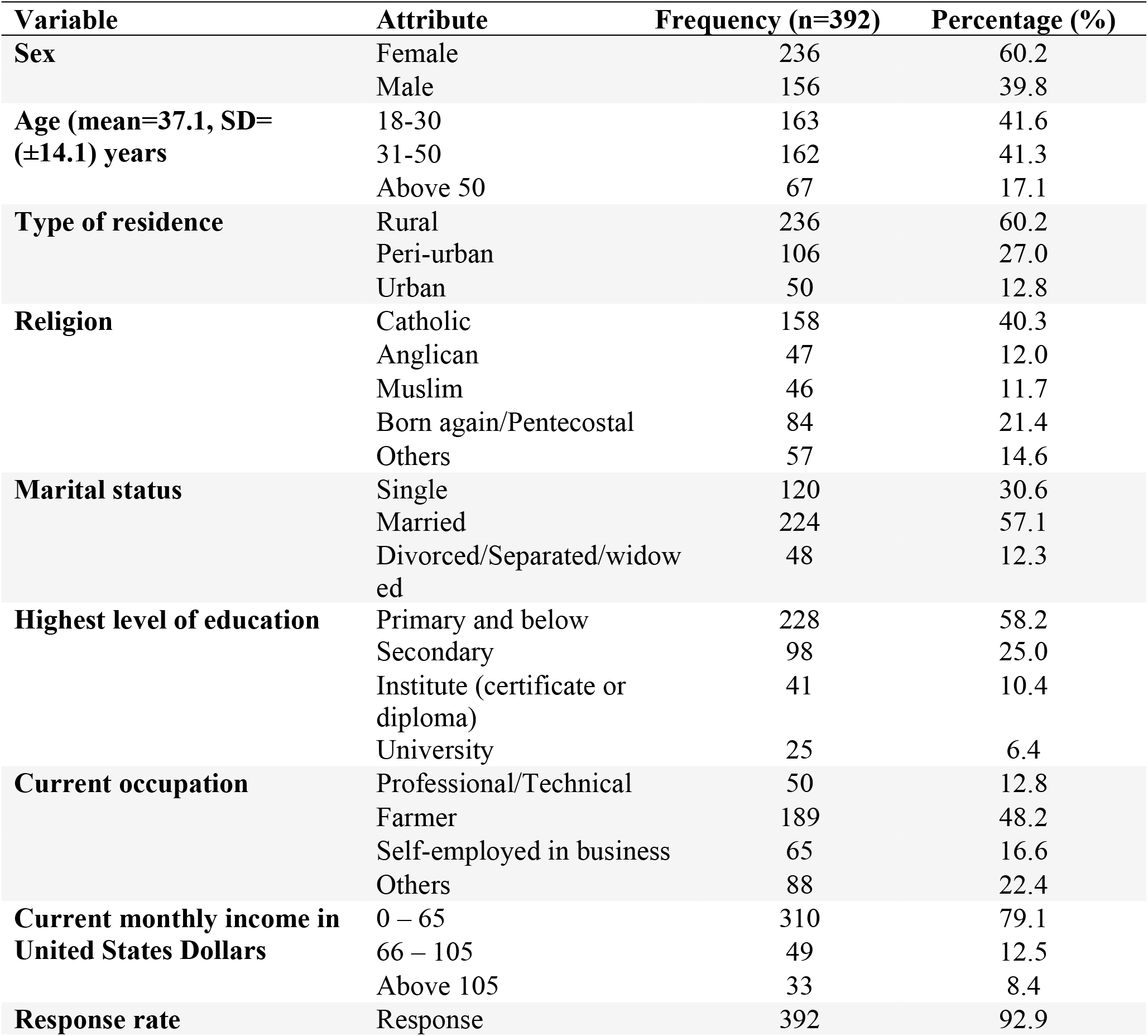
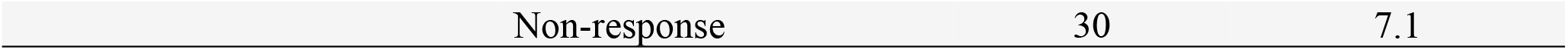
Socio-demographic characteristics of adults PLHIV in Kamuli district.

| Variable | Attribute | Frequency (n=392) | Percentage (%) |
| --- | --- | --- | --- |
| **Sex** | Female | 236 | 60.2 |
|  | Male | 156 | 39.8 |
| **Age (mean=37.1, SD= (±14.1) years** | 18-30 | 163 | 41.6 |
|  | 31-50 | 162 | 41.3 |
|  | Above 50 | 67 | 17.1 |
| **Type of residence** | Rural | 236 | 60.2 |
|  | Peri-urban | 106 | 27.0 |
|  | Urban | 50 | 12.8 |
| **Religion** | Catholic | 158 | 40.3 |
|  | Anglican | 47 | 12.0 |
|  | Muslim | 46 | 11.7 |
|  | Born again/Pentecostal | 84 | 21.4 |
|  | Others | 57 | 14.6 |
| **Marital status** | Single | 120 | 30.6 |
|  | Married | 224 | 57.1 |
|  | Divorced/Separated/widowed | 48 | 12.3 |
| **Highest level of education** | Primary and below | 228 | 58.2 |
|  | Secondary | 98 | 25.0 |
|  | Institute (certificate or diploma) | 41 | 10.4 |
|  | University | 25 | 6.4 |
| **Current occupation** | Professional/Technical | 50 | 12.8 |
|  | Farmer | 189 | 48.2 |
|  | Self-employed in business | 65 | 16.6 |
|  | Others | 88 | 22.4 |
| **Current monthly income in United States Dollars** | 0 – 65 | 310 | 79.1 |
|  | 66 – 105 | 49 | 12.5 |
|  | Above 105 | 33 | 8.4 |
| **Response rate** | Response | 392 | 92.9 |

### Level of risk perception for hypertension among adult PLHIV in Kamuli district (n=392)

Overall, 68.1% (267/392) of the respondents had a low-risk perception for HTN (Fig 1). Particularly, 62.5% (245/392) believed that their absolute risk of developing HTN in a lifetime was low, and only 31.9% (125/392) believed they had very high likelihood of developing HTN when aged 35+ years Figure 1.

**Figure 1:**
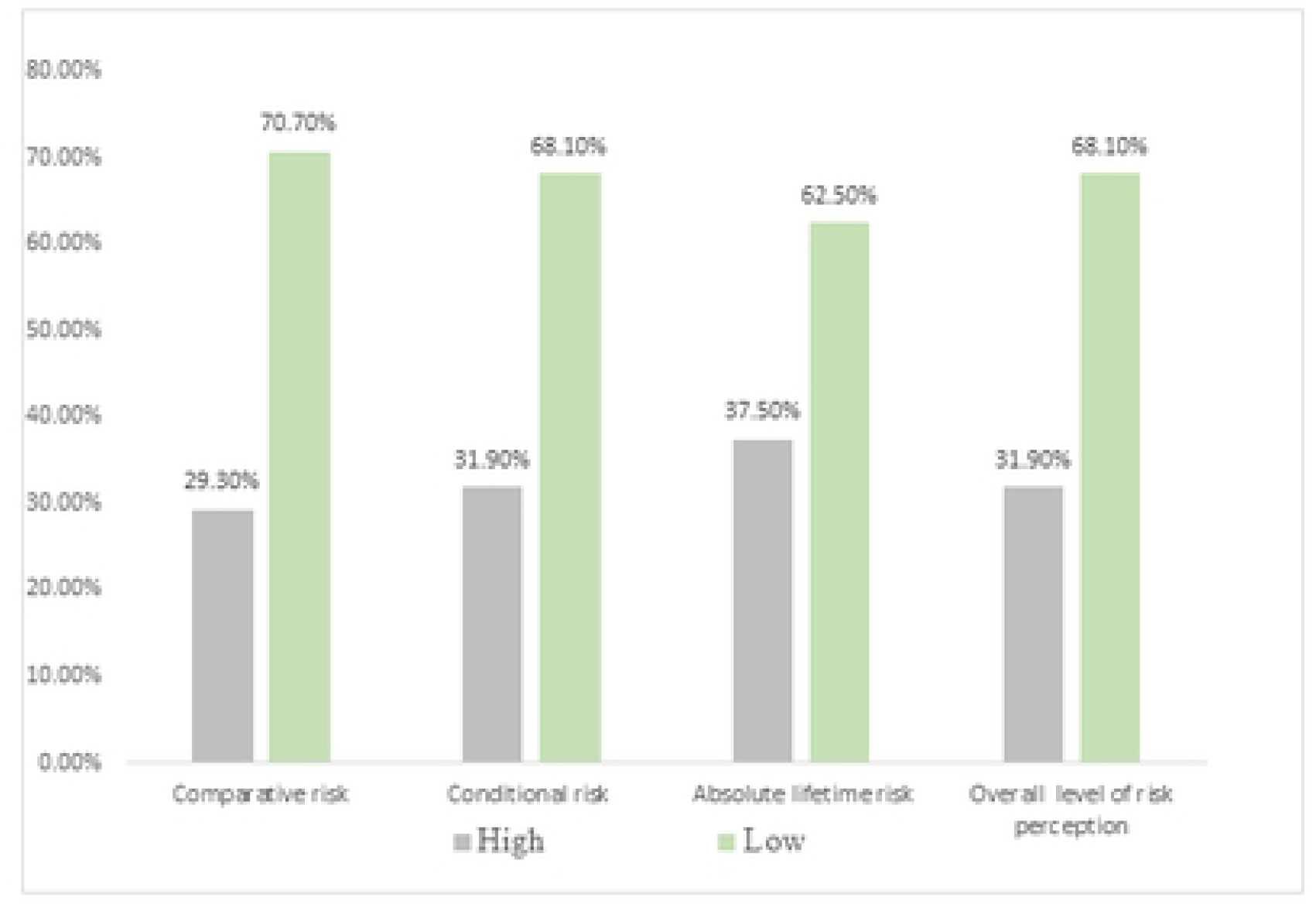
Level of risk perception for HTN among adult PLHIV in Kamuli District, Uganda

### Factors associated with the low level of risk perception for hypertension among adult PLHIV in Kamuli District (n=392)

At bivariate analysis, residing in an urban area was negatively associated with low-risk perception for HTN. On the other hand, having secondary education, low knowledge, low level of trust, having no other NCDs and no family history of HTN were positively associated with low-risk perception for HTN. At multivariable analysis, residence in peri-urban area was positively associated with low-risk perception for hypertension (APR=1.23; 95%CI: 1.04-1.46), while residing in an urban area was negatively associated with a low-risk perception. Compared to those who did not have any formal education, respondents who perceived themselves to be at low-risk among those with secondary level education was 23% (APR=1.23; 95%CI: 1.02 −1.47) and 95% among those with tertiary education (APR=1.95; 95%CI: 1.43-2.67). Furthermore, respondents with a low level of HTN knowledge had a 98% low level of risk perception compared to those with a high level of HTN knowledge (APR=1.98; 95%CI: 1.55-2.53). Additionally, low level of trust for health workers who conduct HTN awareness (APR=1.13; 95%CI: 1.01-1.25), having no other NCD (APR=1.39; 95%CI: 1.01-1.89), having no family history of HTN (APR=1.39; 95%CI: 1.14-1.58), engagement in inadequate vigorous intensity PA (APR=1.23; 95%CI: 1.10-1.37), and consumption of sufficient fruits and vegetables (APR=1.13; 95%CI: 1.01-1.27) were associated with low-risk perception for HTN as shown in Table 2.

**Table 2:** Factors associated with low-risk perception for HTN among adult PLHIV in Kamuli District.

| Variable | Level of risk perception |  | Unadjusted PR<br>at 95% CI | P-values | Adjusted PR at<br>95% CI | P-values |
| --- | --- | --- | --- | --- | --- | --- |
|  | Low<br>(N=267)<br>F (%) | High (N=125)<br>F (%) |  |  |  |  |
| **Age** |  |  |  |  |  |  |
| 18-30 | 127 (47.6) | 36 (28.8) | 1 |  | 1 |  |
| 31-50 | 107 (40.1) | 55 (44.0) | 0.85 (0.74-0.97) | **0.019** | 1.12 (0.99-1.27) | 0.069 |
| 51+ | 33 (12.4) | 34 (27.2) | 0.63 (0.49-0.82) | **<0.001** | 0.73 (0.67-1.03) | 0.096 |
| **Sex** |  |  |  |  |  |  |
| Male | 105 (39.3) | 51 (40.8) | 1 |  |  |  |
| Female | 162 (60.7) | 74 (59.2) | 1.02 (0.89-1.17) | 0.782 |  |  |
| **Residence** |  |  |  |  |  |  |
| Rural | 172 (64.4) | 65 (52.0) | 1 |  | 1 |  |
| Peri-urban | 82 (30.7) | 23 (18.4) | 1.08 (0.95-1.22) | 0.262 | 1.23 (1.04-1.45) | **0.014** |
| Urban | 13 (4.9) | 37 (29.6) | 0.36 (0.22-0.58) | **<0.001** | 0.73 (0.54-0.99) | **0.049** |
| **Education level** |  |  |  |  |  |  |
| None | 29 (10.9) | 19 (15.2) | 1 |  | 1 |  |
| Primary | 114 (42.7) | 66 (52.8) | 1.05 (0.81-1.35) | 0.717 | 1.09 (0.90-1.32) | 0.379 |
| Secondary | 90 (33.7) | 8 (6.4) | 1.52 (1.20-1.93) | **0.001\*** | 1.23 (1.02-1.47) | **0.028\*** |
| Tertiary | 34 (12.7) | 32 (25.6) | 0.85 (0.61-1.18) | 0.341 | 1.95 (1.43-2.67) | **<0.001\*** |
| **Level of HTN knowledge** |  |  |  |  |  |  |
| High | 36 (13.5) | 97 (77.6) | 1 |  | 1 |  |
| Low | 231 (86.5) | 28 (22.4) | 3.29 (2.48-4.37) | **<0.001\*** | 1.98 (1.55-2.53) | **<0.001\*** |
| **Trust in health workers** |  |  |  |  |  |  |
| High | 211 (79.0) | 125 (100.0) | 1 |  | 1 |  |
| Low | 56 (21.0) | 0 (0.0) | 1.59 (1.47-1.73) | **<0.001\*** | 1.13 (1.01-1.25) | **0.025\*** |
| **NCD comorbidity** |  |  |  |  |  |  |
| Yes | 20 (7.5) | 67 (53.6) | 1 |  | 1 |  |
| No | 247 (92.5) | 58 (46.4) | 3.52 (2.39-5.20) | **<0.001\*** | 1.38 (1.00-1.89) | **0.049\*** |
| **Family history of HTN** |  |  |  |  |  |  |
| Yes | 73 (27.3) | 111 (88.8) | 1 |  | 1 |  |
| No | 194 (72.7) | 14 (11.2) | 2.35 (1.96-2.82) | **<0.001** | 1.42 (1.20-1.68) | **<0.001\*** |
| **Vigorous intensity PA** |  |  |  |  |  |  |
| Adequate | 164 (61.4) | 80 (64.0) | 1 |  | 1 |  |
| Inadequate | 103 (38.6) | 45 (36.0) | 1.04 (0.90-1.19) | 0.621 | 1.23 (1.10-1.37) | **<0.001\*** |
| **Currently smoker** |  |  |  |  |  |  |
| Yes | 7 (2.6) | 9 (7.2) | 1 |  |  |  |
| No | 260 (97.4) | 116 (92.8) | 1.58 (0.90-2.77) |  |  |  |
| **Fruit/vegetables intake** |  |  |  |  |  |  |
| Insufficient | 166 (62.2) | 119 (95.2) | 1 |  | 1 |  |
| Sufficient | 101 (37.8) | 6 (4.8) | 1.62 (1.45-1.81) | **<0.001\*** | 1.13 (1.01-1.27) | **0.047\*** |

## Discussion

This study assessed the level of risk perception for HTN and the associated factors among adult PLHIV in Kamuli District. More than half of the respondents perceived to be at low-risk for HTN and associated factors were: having higher education level, low HTN knowledge, peri-urban residency, having low trust in health workers, having no family history of HTN and no NCD comorbidities.

Although high-risk perception is essential for an individual to take timely and efficient behavioural measures to prevent development of a given disease or health condition, our study found that 68.1% of the respondents had a low-risk perception for HTN. The low-risk perception may be attributed to the observed higher proportion of respondents with low knowledge about HTN, low prevalence of NCD comorbidities, and low proportions of respondents with family history of HTN. Similarly, low-risk perception for HTN was reported in Kenya with the majority of the respondents having inappropriate attitude towards the preventive measures for HTN [22]. Related findings were reported by a study conducted among adults with HTN in Ethiopia where majority (58.6%) of the participants had a low-risk perception for CVDs [23]. Our findings were also consistent with a study conducted among adult PLHIV in Rwanda which reported that respondents had low risk perception for NCDs [24]. The observed low-risk perception for HTN highlights the need for targeted interventions to improve HTN risk perception and promote uptake of preventive behaviours among PLHIV.

Our study found that residing in a peri-urban area was positively associated with low-risk perception for HTN, which may be due to limited information (thus low knowledge) among PLHIV in peri urban areas thus perceive low risk. In contrast, respondents who resided in urban areas perceived to be at higher risk for HTN compared to rural dwellers, possibly due to easy access to health information on Televisions and radios. An informed population is more likely to perceive higher risk and make healthy lifestyle choices and adopt positive health-oriented behaviours [25]. Surprisingly, we found that respondents who had attained higher education levels (secondary and tertiary) were more likely to have low-risk perception for HTN compared to those with no education. People with a formal education may have higher knowledge about HTN and thus adopt and maintain its preventive measures and may perceive being at low risk. Relatedly, among people living with HIV in Chitungwiza Hospital Zimbabwe, having low education level (primary or less) was associated with negative perception about NCD management compared to those with higher education (P<0.01) [26]. On the otherhand, a study by Cioe et al. [27] found that knowledge of HTN risk factors did not predict perceived risk for HTN among PLHIV in Rhode Island Hospital -USA (p>0.05). Contrary to findings of the USA study, our study findings indicated that respondents with low knowledge of HTN had low risk perception for the condition compared to those with high level of knowledge. These findings underscore the need for health education interventions to improve HTN knowledge and facilitate reduction of engagement in behavioural risks for HTN. These education interventions need to be tailored to the educational levels of the target population and integrated into HIV care services. For our study, age did not independently predict risk perception for HTN, which contract with findings of a study conducted among adult men in Nigeria, where respondents’ perceived risk for prostate cancer increased with age (β=0.014) [28]. Relatedly, a cross sectional survey among South African adult men found that age of respondent independently predicted level of risk perception for NCDs [14]. Our study found that respondents with high level of trust in health workers who conduct HTN awareness had a higher risk perception for HTN compared to those who had low level of trust. Having high trust in health workers may increase the patients’ HTN risk perception due to higher confidence in the health information. Similar to our findings, trust has been reported to play a significant role in predicting people’s health risk perceptions and disease preventive behaviors in Uganda [29]. Relatedly, client trust in CHWs offering maternal and newborn health education was reported to result into acceptability of the health services and responsiveness to health education programs [30]. Our findings suggest that the more people trust the health workers and the risk awareness information received, the more likely they will accept and believe that they are at higher risk for that particular condition. Our findings highlight the need for strengthening healthcare provider training on the link between HIV and HTN to enhance their confidence in HTN risk communication to PLHIV and improve their trust by PLHIV. Besides, we found that respondents with no NCD comorbidities perceived themselves to be at low-risk for HTN compared to those with comorbidities. People without NCD comorbidities are likely to self-rate their general health status as good hence perceive low-risk for HTN. Besides, people with one or more NCDs are more likely to seek for more information on risk factors and thus gain much knowledge which can influence them to perceive higher risk for other NCDs. Individuals with no underlying NCDs may feel not bothered about HTN thereby perceiving to be at low risk. Our study also found that respondents without a family history of HTN had a low-risk perception for HTN compared to those with one. This could be because PLHIV with a family history are more likely to consider themselves susceptible and believe they may develop the disease. While local context literature on the association between HTN risk perception and family history is limited, studies elsewhere have reported a positive association between family history of HTN and perception of risk for CVDs (p<0.01) [31,32]. Indeed, there is available literature which suggests that genetic factors are responsible for about 30% change in blood pressure [33]. Our findings were in agreement with a study conducted among female students in Ghana where respondents without a family history of breast cancer were 90% less likely to perceive breast cancer risk (AOR = 0.10, 95% CI = 0.04– 0.29) compared to those with family history of the cancer [34]. Relatedly, adult persons in Johannesburg (South Africa) who had a family medical history of NCDs self-perceived to be at high-risk of developing NCDs [14]. These findings reveal the need to develop tailored risk awareness interventions for PLHIV with neither NCD comorbidities nor family history of HTNs so as to increase their level of HTN risk perceptions.

### Study strengths and limitations

We believe that there were key strengths in our findings; question items used to measure risk perception (primary outcome) were adapted from a tool validated through a recent study conducted among adult women in Uganda [17]. This increased internal validity of the findings. However, these findings are subject to limitations. For instance, purposive inclusion of high-volume health facilities and the consecutive sampling used for this study are prone to selection bias. Besides, the self-reporting method used to measure some of the variables is subject to recall bias. Nonetheless, our study considered a fairly large sample size to increase validity.

## Conclusion

This study found that that a higher proportion of PLHIV (68.1%) perceived to be at low-risk for HTN, despite the fact that PLHIV are generally a high-risk group given the relationship between HIV and HTN/NCDs. Key factors independently associated with low-risk perception for HTN were having low HTN knowledge, low trust in health workers, having no family history of HTN and no NCD comorbidity. There is need to implement appropriate health education programs specifically tailored to increasing HTN knowledge and risk perception among PLHIV. Besides, there is need to develop and implement risk awareness interventions for PLHIV with neither NCD comorbidities nor family history of HTNs to increase their level of HTN risk perception. Health workers should integrate routine HTN risk assessments into HIV care services to identify PLHIV who are at very high-risk of developing HTN and provide them with personalized care. By implementing these strategies, it is possible to improve knowledge and risk perception for HTN and foster uptake of preventive behaviours but also promote early detection and management of HTN among adult PLHIV.

## Data Availability

We have attached the utilized data set with no participant identity

## Supplementary information

1. Figure 1. : Level of risk perception for HTN among adult PLHIV in Kamuli District, Uganda
2. Data set in excel

## Notes

### Competing Interest Statement

The authors have declared no competing interest.

### Funding Statement

Authors received no funding for this research work

### Author Declarations

Makerere University Higher Degrees Research and Ethics Committee

